# Self-Care, Health Literacy and Their Associations Amongst Patients with Chronic Kidney Disease (CKD) in Primary Care

**DOI:** 10.1101/2022.03.02.22271758

**Authors:** Han-Kwee Ho, Eileen Yi-Ling Koh, Wan-Ching Chua, Meykkumar S/O Meyappan, Ray Ern Chung, Pereira Emma Marie-Pamerlyn, Chenlu Zhai, Leon Jian-Ying Lim, Adina Abdullah, Ngiap-Chuan Tan

## Abstract

**Aim:** The study objective was to determine the levels of self-care (including domains of behaviour, motivation, self-efficacy) and health literacy, and study their associations amongst patients with chronic kidney disease (CKD) in primary care setting in Singapore.

**Method:** A cross-sectional, questionnaire-based study was conducted in one public-sector primary care clinic. Participants aged 21 to 80 years with hypertension were recruited from the clinic CKD register with 5,500 patients. Self-care profile (including behaviour, motivation and self-efficacy) were measured using Hypertension Self-Care Profile (HTN-SCP, range 0-240, domain range 0-80). Health literacy was measured using Short-Form Health Literacy Scale (HLS-SF12, range 0-50, limited literacy ≤33).

**Results:** A total of 347 out of 354 randomly selected patients consented to participate in the study. Two hundred and eighty-nine fully-completed responses were analysed. The mean self-care (HTN-SCP) score was 182.7 (SD 23.2), while mean scores were 55.3 (SD 8.6), 63.3 (SD 8.7), 64.0 (SD 9.3), for behaviour, self-efficacy and motivation domains respectively. The mean health literacy score was 36.1 (SD 7.7), and 31.1% of participants had limited health literacy. Limited health literacy was associated with self-efficacy (OR= −7.2, 95%CI=−9.1 to −5.2, p<0.001), motivation (OR= −6.1, 95%CI=−8.3 to −3.9, p<0.001) and behaviour (OR= −4.5, 95%CI=−6.6 to −2.4, p<0.001). Self-care was not associated with age, CKD status, household income and education but was associated with gender and limited health literacy. In the final regression model only limited health literacy was associated with self-care scores (Adjusted beta −17.4, p<0.001).

**Conclusion:** One-third of the patients with CKD in primary care had limited health literacy. Self-care was not associated with age, gender, CKD status, household income or education. Limited health literacy was associated with self-care, with strongest association with self-efficacy, followed by motivation and behaviour. More targeted approach can be adopted to improve self-care and health literacy amongst patients with CKD.

## Introduction

Chronic kidney disease (CKD) is associated with significant health burden not only because of the impact of the disease but also because of the economic impact to the patient and health system. CKD accounted for a global prevalence of 13.4%^1^ in 2016 and a 41.5% increase in global all-age mortality rate attributable to CKD between 1990 and 2017^2^. Globally, CKD due to diabetes and hypertension showed a 107% and 28% increase in age-standardised death rate respectively^3^. In Singapore, CKD ranked 7th as cause of Years of Life lost^4^, with Crude Incidence Rate (CIR) increasing from 383.9 per million population (pmp) in 2010 to 503.0 pmp in 2018^4^. In fact, the current CKD prevalence of 12% is projected to double to 24% by 2035^5^.

In 2011, a CKD intervention programme^6^ aimed at identifying patients with early CKD and optimising renal-protective therapy was launched^7^ in Singapore. This programme was rolled out by the local Ministry of Health as the HALT-CKD^8^ (Holistic Approach in Lowering and Tracking Chronic Kidney Disease) programme in 2017 to remind and encourage primary care physicians in public sector to prescribe renal-protective medicine such as Angiotensin-Converting Enzyme Inhibitors (ACEi) and Angiotensin-Receptor Blockers (ARBs) to patients when clinically indicated^9^. Under this programme, approximately 90% of patients with CKD have already been placed on these renal-protective medicine^10^.

However, more can be done to increase patient involvement to retard the progression of CKD. A recent local qualitative study^11^ found that a lack of awareness of CKD, knowledge of the disease and a passive attitude towards self-management was a major barrier to CKD self-management. Patient self-management behaviour includes lifestyle modifications, compliance to medication, follow-up and self-monitoring of parameters like blood pressure. For patients with CKD, this often means reduced salt in diet and keeping both blood pressure and glucose level (for those with diabetes) within treatment goals. These behaviours have been shown to slow CKD progression^12,13,14^ and reduce its morbidity and mortality^15,16,17^.

Low health literacy has been identified as a key barrier to development of self-management skills^18^, and conversely high health literacy often translates to better patient self-care, patient education and engagement^19,20^. While health literacy has been known to be variable in Asian population^21,22^, no health literacy data related to CKD is currently available for Singapore. Patients with higher health literacy may have more confidence (self-efficacy) in their abilities and positively influence their self-management behaviours^23,24^. Self-efficacy refers to the person’s belief (or confidence) in their capacity to carry out the behaviour to achieve the outcome. Self-efficacy and motivation have also been independently studied to be closely associated with self-management behaviour^25,26,27,28^ and are also pivotal in facilitating adherence and lifestyle changes among patients^28,29^.

The aim of the study is to determine the levels of self-care (including behaviour, self-efficacy, motivation) and health literacy, and their associated factors in primary care patients with CKD.

## Method

### Study Design

This was a cross-sectional, questionnaire-based study conducted in one of the 20 public sector primary care clinics in Singapore. The clinic is located in the eastern part of Singapore (Pasir Ris Town) and sees more than 600 patients a day with an average of 14 attending primary care physicians and a total staff strength of about 110. The clinic sees a mixture of both chronic follow-up visits as well as acute conditions. A separate section for patients with fever or upper respiratory tract symptoms was set up since early 2020 as part of COVID-19 response. This section of the clinic was excluded from the study.

### Study population

Adult patients above age 21 and below 80-year-old who have essential hypertension, have CKD stage 1 to 5 and who were being followed up at the clinic were included in the study. The upper age limit was chosen to align with the Singapore Ministry of Health HALT-CKD programme. The key study interest was to study CKD secondary to chronic diseases. Thus, patients with nephrotic syndrome, other forms of glomerulonephritis or renal parenchyma diseases were excluded. Patients who were known to have dementia, either from attending physician’s notes or self-reported, and those with any other disabilities or conditions that prevent them from completing the questionnaires were also excluded.

### Study Instruments

#### Hypertension Self-Care Profile (HTN-SCP)

For self-management behaviour, motivation and self-efficacy, the Hypertension self-care profile (HTN-SCP)^30^ was used. Diabetes and hypertension accounted for the vast majority of End-Stage Renal Failure (ESRF) in Singapore^31,32^. There may be some limitations in using a self-care tool for hypertension, versus a self-care tool specific for CKD^33,34,35^. Using a CKD-specific self-care tool was considered, but eventually a locally validated tool with a comprehensive coverage of self-care domains was deemed more appropriate. The HTN-SCP has been validated in Singapore^36^, in the same public sector primary care clinic setting. Public and private primary care services run in parallel in Singapore, with the public sector receiving more government subsidy and possibly seeing more patients of higher complexity. HTN-SCP covered multiple self-care domains such as medication adherence, physical activity, diet restriction, alcohol consumption, smoking, home blood pressure monitoring, weight control, regular follow-up, and stress management. These domains were often common across cardiovascular related chronic diseases including CKD.

The internal consistency for this instrument is good, with Cronbach’s alpha coefficients being 0.83 for behaviour, 0.93 for motivation and 0.91 for self-efficacy. The HTN-SCP showed high construct validity, and there was no floor or ceiling effect for the 3 subscales. This tool measures all 3 variables using the same set of 20 questions using different opening instructions: from “How often do you do the following” for self-management behaviour to “How important is it for you to do the following?” for motivation and “How confident are you that you could do the following?” for self-efficacy. For each section, the minimum score is 0 and the maximum score is 80, with a total maximum self-care score of 240. Higher scores indicate higher level of self-care, behaviour, motivation or self-efficacy.

#### Short-Form Health Literacy Instrument (HLS-SF12)

The HLS-SF12 was used for the assessment of health literacy. This instrument was derived from the 47-item European Health Literacy Questionnaire (HLS-EU-Q47)^37^ and has been validated in 6 Asian countries including Malaysia and Taiwan^38^. The HLS-SF12 was demonstrated to have high reliability with a Cronbach’s alpha of 0.85 (range of 0.79-0.90, with Malaysia and Taiwan both 0.85). There was also good criterion-related validity, a moderate and high level of item-scale convergent validity, and no floor or ceiling effect throughout the populations in the six countries. The health literacy score can range from 0 to 50, with higher scores indicating higher health literacy. A score of 33 and below indicates limited literacy. The 12 questions also measured the 3 domains^37^ of health literacy, namely health care, disease prevention and health promotion.

#### Sample size

Primary and secondary outcomes of self-care (including behaviour, motivation, self-efficacy) and health literacy were factors considered when considering the appropriate sample size. Based on Koh et al^36^, the mean and standard deviation of self-care score was 181.8 and 28.3 respectively. Assuming that the sample has high self-care score of 192 with a 95% confidence level^39^, the minimum sample size required was 63. However, a larger sample size was required when health literacy was considered. Abd-Rahim et al^40^ reported a mean proportion of those who have adequate health literacy as 80% (334/413). Using sample size for proportion, with a total population of 5500 CKD patients in clinic CKD register, the minimum required sample is 229, with 95% confidence interval and 5% precision. With a buffer of 20% to account for incomplete data, we aim to recruit at least 290 to cover both outcomes.

#### Sampling method

A hybrid sampling method was adopted to complete this study within a reasonable time frame in the context of resource constraints and revised research protocols due to safe-distancing policies. A list of appointments for patients with CKD in the clinic was generated every week in chronological order based on room numbers assigned to patients and appointment times. Patients were numbered in a chronological order based on this list every week. A random number generator was used to identify the patient to be invited to participate in the study. The number of random numbers generated was dependent on the research staff available for the day. A new random number was used if the selected patient declined participation or has been excluded.

The attending physician was informed on the day of patient’s appointment. The attending physician would first screen for any of the exclusion criteria as aforementioned. If found to be suitable, the attending physician would then verbally invite the identified patient to participate in the study. Patients who agreed to participate would be directed to a separate room where trained research assistants would obtain informed consent and administer the questionnaires. Participants would be given a S$10 (about US$7.40) supermarket voucher on completion of the questionnaire.

This study has been approved by ethics committee of SingHealth Centralised Institution Review Board (CIRB Reference 2020/3109).

#### Statistical Analysis

Descriptive statistics were summarized using frequencies and percentages. Hypertension Self-care Profile (HTN-SCP) total scores and their individual domain scores (behaviour, motivation, self-efficacy) were analysed with participant demographics using independent t-test and the ANOVA test. Linear regression was used to explore the relationship between the self-management behaviour with health literacy, self-efficacy and motivation.

## Results

A total of 594 patients were screened and 222 declined participations. A further 18 who were screened were not selected as they did not fulfil inclusion criteria. Of the remaining 354 who consented, 4 did not proceed to complete the survey, while 2 declined participation subsequently. One participant was excluded due to error in her age computation. Out of the remaining 347 participants, 58 were excluded due to missing data in the questionnaire (**Figure 1**). Eventually, complete data of 289 participants were analysed. **Table 1** shows the demographics of participants and their levels of self-care score, behaviour, motivation, self-efficacy and health literacy.

**Table 1.**
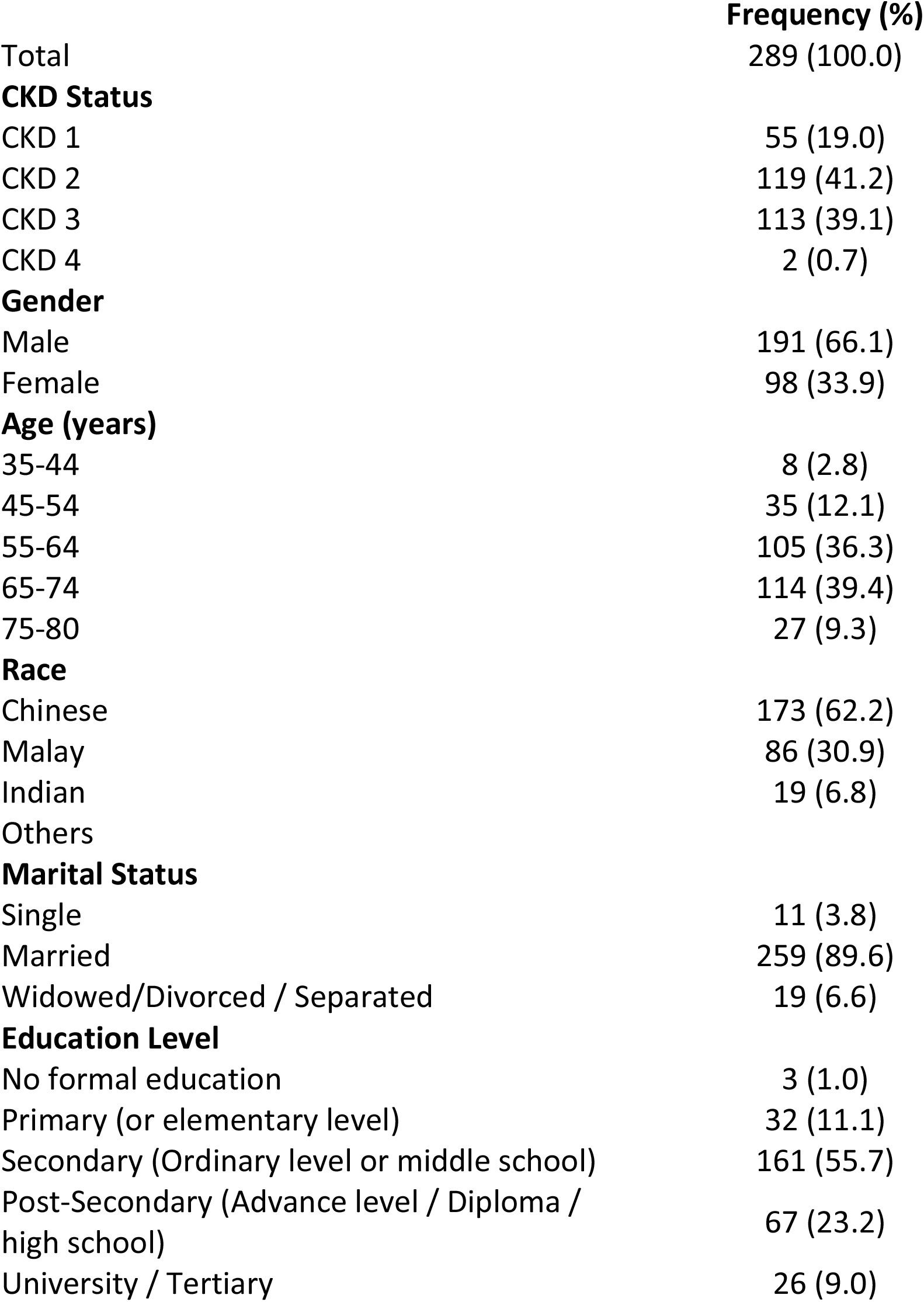
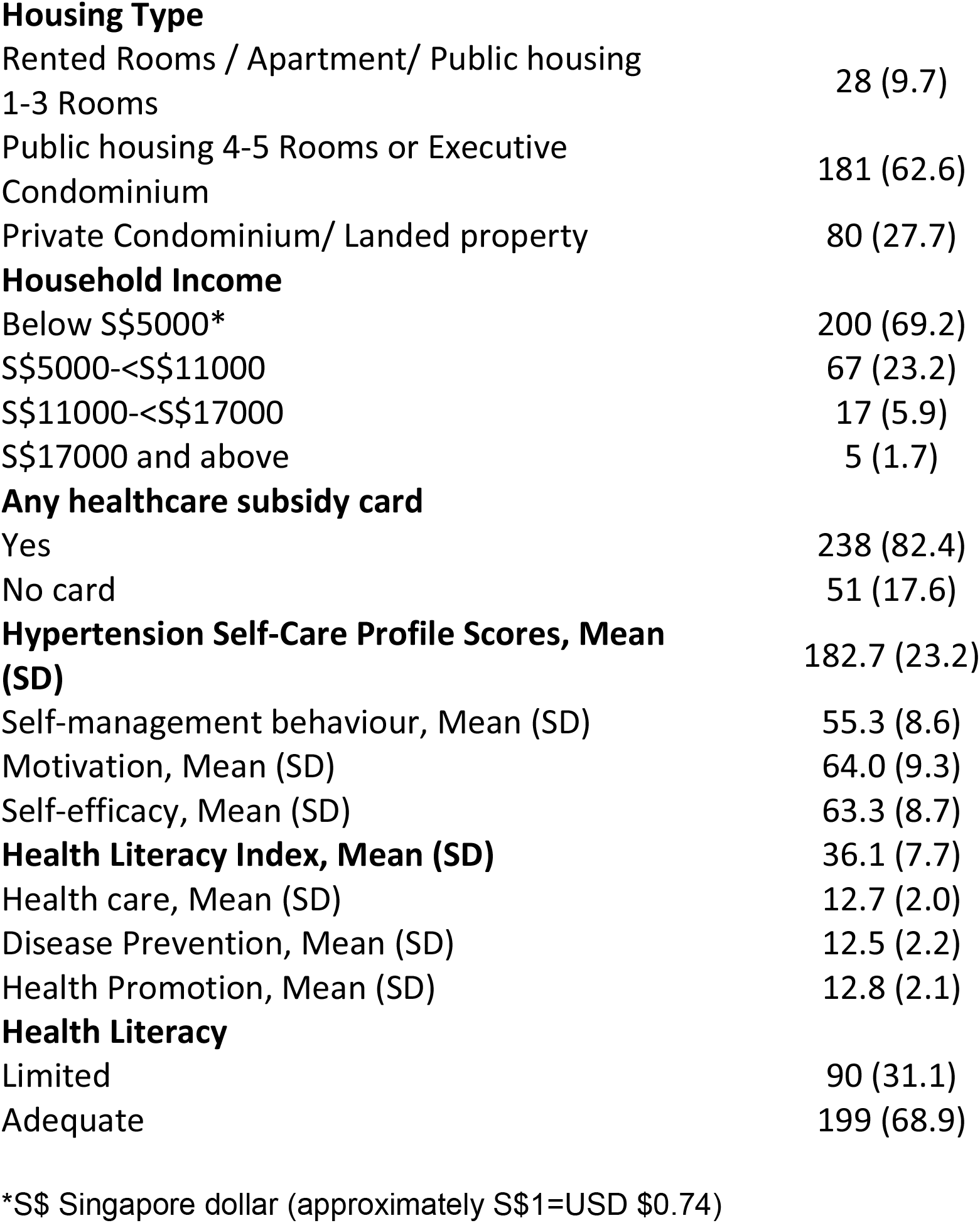
Demographics of Participants and Their Levels of Hypertension Self-Care Profile (HTN-SCP) scores, Behaviour, Motivation, Self-Efficacy and Health Literacy

**Figure 1:**
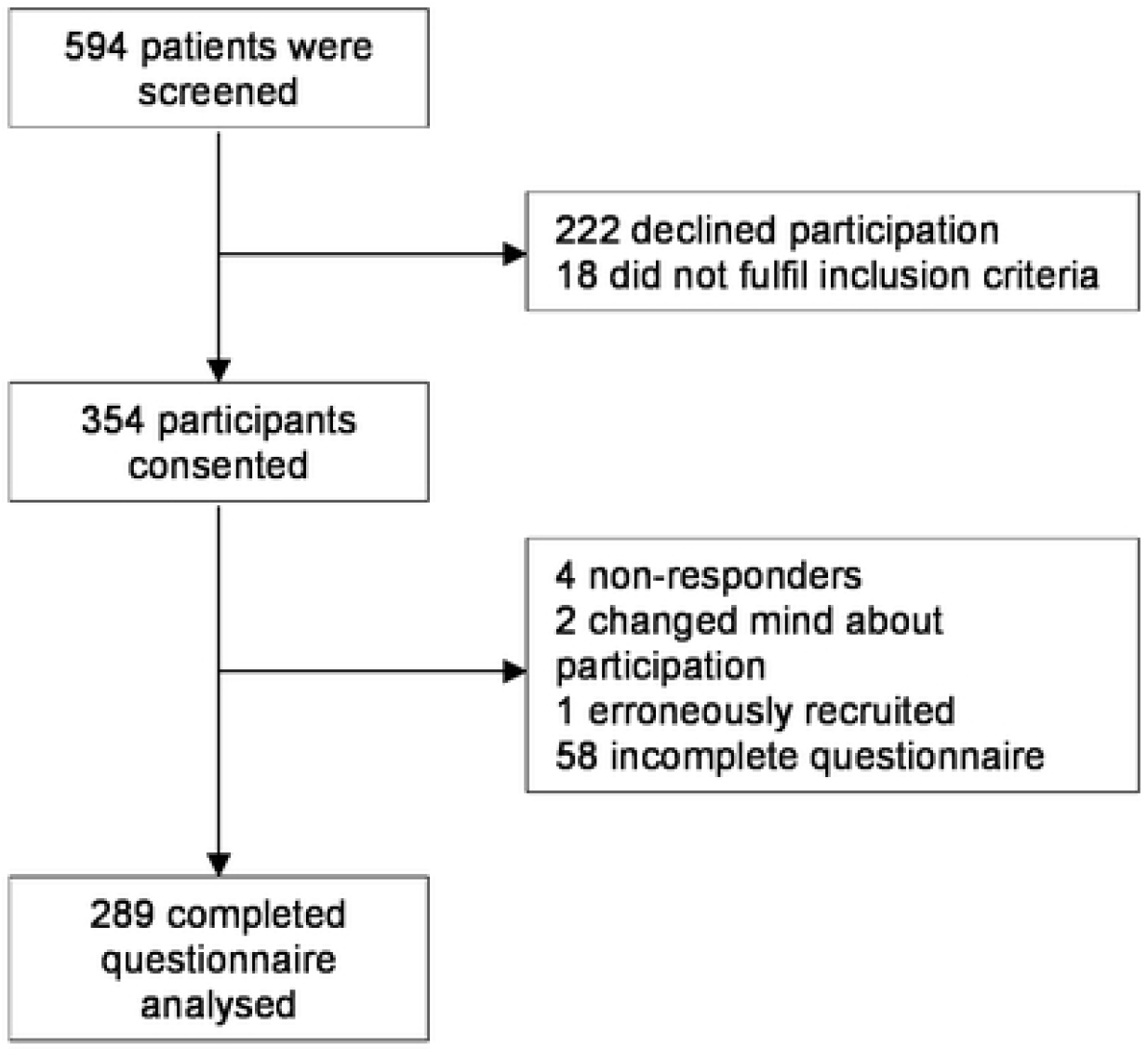
Participant’s recruitment flow.

The average age of the participants was 63.7 years old (SD 8.7, range 35 to 79). A total of 66.1% (n=191) of participants were male, with 62.2% (n=173) Chinese, 30.9% (n=86) Malays and 6.8% 9 (n=19) Indians. The bulk of the participants were at CKD stage 2 and 3 (80.3%), with none at stage 5 and only 2 were at CKD stage 4. The mean total HTN-SCP self-care score was 182.7 (SD 23.2), with a mean behavioural score of 55.3 (SD 8.6), mean motivation score was 64.0 (SD 9.3) and mean self-efficacy score of 63.3 (SD 8.7).

The mean health literacy score was 36.1 (SD 7.7). The mean scores of the 3 domains of health care, disease prevention and health promotion range from 12.5 to 12.8, with a SD of between 2.0 to 2.2. Some 31.1% had score of 33 and below, fulfilling the criteria for limited health literacy.

### Self-Care Scores

**Table 2** showed the baseline characteristics of study participants and their association with HTN-SCP self-care scores.

**Table 2.** Factors Affecting Self-Care Scores

|  | **Hypertension**<br>**Self-Care Profile**<br>**score** | **p-value** | **Adjusted Beta**<br>**(95% CI)** | **p-value** |
| --- | --- | --- | --- | --- |
| Total | 182.7 (23.2) |  |  |  |
| **CKD Status** |  | 0.767 |  |  |
| CKD 1 | 183.1 (19.4) |  | - | - |
| CKD 2 | 182.9 (22.8) |  | - | - |
| CKD 3 | 181.9 (25.4) |  | - | - |
| CKD 4 | 199.0 (25.5) |  | - | - |
| **Gender** |  | 0.039 |  |  |
| Male | 180.6 (23.8) |  | -4.7 (-10.1-0.7) | 0.087 |
| Female | 186.6 (21.7) |  | Ref | - |
| **Age (years)** |  | 0.497 |  |  |
| 35-44 | 171.5 (16.7) |  | - | - |
| 45-54 | 179.7 (24.2) |  | - | - |
| 55-64 | 184.6 (24.3) |  | - | - |
| 65-74 | 183.0 (22.1) |  | - | - |
| 75-80 | 180.6 (23.8) |  | - | - |
| **Race** |  | 0.104 |  |  |
| Chinese | 180.1 (23.9) |  | -6.6 (-16.9-3.7) | 0.207 |
| Malay | 186.2 (22.0) |  | -2 (-12.8-8.8) | 0.713 |
| Indian | 186.6 (23.9) |  | Ref | - |
| Others |  |  |  |  |
| **Marital Status** |  | 0.465 |  |  |
| Single | 174.2 (19.7) |  | - | - |
| Married | 182.9 (23.1) |  | - | - |
| Widowed/Divorced /<br>Separated | 183.7 (26.4) |  | - | - |
| **Education Level** |  | 0.874 |  |  |
| No formal education | 183.0 (10.1) |  | - | - |
| Primary (or elementary<br>level) | 183.4 (20.2) |  | - | - |
| Secondary (Ordinary level or middle school) | 181.4 (24.2) |  | - | - |
| Post-Secondary (Advance level / Diploma / high school) | 183.6 (24.0) |  | - | - |
| University / Tertiary | 186.2 (20.7) |  |  |  |
| **Housing Type** |  | 0.399 |  |  |
| Rented Rooms / Apartment/ Public housing 1-3 Rooms | 187.5 (22.1) |  | - | - |
| Public housing 4-5 Rooms or Executive Condominium | 181.5 (23.1) |  | - | - |
| Private Condominium/ Landed property | 183.6 (23.9) |  | - | - |
| **Household Income** |  | 0.486 |  |  |
| Below S\$5000* | 183.9 (22.8) | | - | - |
| S\$5000-<S\$11000 | 179.0 (25.3) | | - | - |
| S\$11000-<S\$17000 | 181.6 (21.5) | | - | - |
| S\$17000 and above | 187.0 (17.6) | | - | - |
| **Any subsidy card** |  | 0.477 |  |  |
| Yes | 183.1 (22.9) |  | - | - |
| No card | 180.5 (24.8) |  | - | - |
| **Health Literacy** |  | <0.001 |  |  |
| Limited | 170.4 (24.4) |  | -17.4 (-22.8--11.9) | <0.001 |
| Adequate | 188.2 (20.5) |  | Ref | - |

\*S\$ Singapore dollar (approximately S\$1=USD \$0.74)

CKD disease stages, age, race, education, housing type and income were not significantly associated with self-care scores. Female gender was associated with higher total self-care score (mean=186.6, SD 21.7, compared with male, mean=180.6, SD 23.8, p=0.039). Limited health literacy was associated with lower self-care scores (mean=170.4, SD 24.4, compared with adequate health literacy, mean 188.2, SD 20.5, p<0.001). However, linear regression analysis showed that only limited health literacy was associated with lower self-care scores (adjusted Beta −17.4, 95% CI −22.8 to −11.9, p<0.001).

**Table 3** shows the result of linear regression of health literacy, motivation and self-efficacy with behaviour. Limited health literacy was associated with self-efficacy (OR=−7.2, 95%CI=−9.1 to −5.2, p<0.001), motivation (OR=−6.1, 95%CI=−8.3 to −3.9), p<0.001) and behaviour (OR=−4.5, 95%CI=−6.6 to −2.4, p<0.001).

**Table 3.**
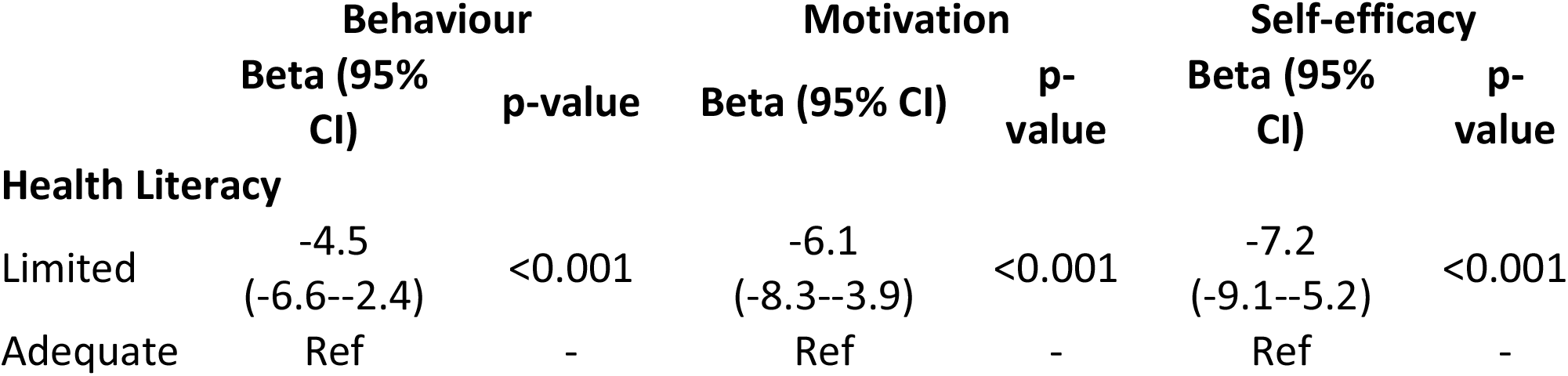
Linear Regression of Health literacy on outcome

|  | Behaviour |  | Motivation |  | Self-efficacy |  |
| --- | --- | --- | --- | --- | --- | --- |
|  | Beta (95% CI) | p-value | Beta (95% CI) | p-value | Beta (95% CI) | p-value |
| **Health Literacy** |  |  |  |  |  |  |
| Limited | -4.5<br>(-6.6--2.4) | <0.001 | -6.1<br>(-8.3--3.9) | <0.001 | -7.2<br>(-9.1--5.2) | <0.001 |
| Adequate | Ref | - | Ref | - | Ref | - |

## Discussion

This study reports the levels of self-care and health literacy amongst participants with CKD in primary care in Singapore. To the best of our knowledge, this was the first time such data has been reported.

It would be difficult to compare self-care levels across populations without considering various factors like measurement instrument used. In another local primary care study by Wee YMS et al^41^ in 2016/17 using the same instrument, the self-care score was reported as 190 (SD 28) while the behaviour domain score was 58 (SD 9). These scores were higher than that found in this study. However, the proportion of participants with CKD and related complications was not reported by Wee YMS et al^41^. In this study, participants have established complications and may have more complex self-care activities to perform. Given that the two studies were conducted in the same setting and measured using the same instrument, the self-care scores for CKD patients were postulated to be lower than that of patients with essential hypertension alone. Another factor that could have played a part was the distribution of the races in the two studies. The proportion of Malays were higher in this study (30.9% in this study, compared to 9.5% in Wee et al) and lower in Indians (6.8% in this study, compared to 19.2% in Wee et al). However, both studies were cross-sectional studies and it would be difficult to establish the temporal relationship of whether lower self-care and behaviour scores amongst participants with hypertension resulted in higher incidence of CKD.

In this study, self-care amongst patients with CKD was not associated with CKD stage, age, income status or education status. In contrast, Tsai YC et al^42^ found that older patients had better self-care behaviour while younger ones had better disease knowledge. The same study reported self-care behaviour increased as renal function deteriorated. The lack of association in this study could be due to the (primary care) setting and participant demographics of this study. Majority of the participants (80.3%) had CKD 2 or 3 in this study while 75.7% were between 55 to 74 years-old. The only significant association with low self-care score from this study was with limited health literacy. This suggest that health literacy is a prerequisite for patients to acquire the relevant knowledge and to apply the acquired knowledge in their own behaviour^43^.

The mean health literacy index in our study was 36.1, even as 87.9% of the participants had secondary (ordinary level or middle school) and above education. The proportion of participants with limited health literacy in this study was almost one-third. Using a different health literacy instrument, Asharani et al^44^ reported a 19.5% limited health literacy in a nationwide survey on diabetes in Singapore. Pooled prevalence of limited health literacy amongst US patients with CKD not on dialysis was 25%^45^, while that for UK was 22.7% (range of 9 to 32%)^46^, whereas that for Taiwan was reported to be 53.4%^43^. This study placed the limited health literacy proportion amongst patients with CKD in Singapore slightly below that of US and UK, while better than that for Taiwan.

The association between health literacy and the 3 domains of self-care namely behaviour, self-efficacy and motivation were consistent with findings in other studies^47^. The association between health literacy and self-efficacy was particularly strong and further expand the association roadmap that may potentially influence self-care positively. One way of looking at improving self-care could be to improve health literacy, aiming to translate improved health literacy to better self-care behaviour. The association between health literacy and self-efficacy, another domain of self-care, could suggest that through improving patient self-confidence, self-care can also improve^48^. Systemic review of evidence suggested that lifestyle modifications targeting at diet and physical activities that were anchored on education *and* modelling of behavioural change through patient confidence-building have been shown to result in significant improvement in outcomes amongst CKD patients^49^.

This study has implications for local primary care practice. Much more can be done in clinical practice to identify the 1-in-3 patients with CKD and limited health literacy to improve their health literacy. In our local setting, each public sector clinic is supported with a CKD coordinator to help in patient education. This service, however, was often underutilised. In addition to patient education, the CKD coordinator could be upskilled with coaching skills to support patient self-confidence and behavioural change.

Beyond physician or clinic level efforts, perhaps a more systematic or programme-based approach towards patients with CKD to improve health literacy could be more effective, complementing system efforts like the HALT-CKD programme to improve clinical outcomes. These efforts could focus beyond behavioural change from improving health literacy but also to work on improving patient self-efficacy and motivation. This would be a more holistic approach to improving self-care for patients with CKD.

This study has its limitations. Other parameters which may affect self-care included perceived health status^50^, anxiety^51^, depression^52^, compliance^53^ and family (or social) support^54,55,56^ have not been examined. Patient well-being and patient activation, defined as patients’ understanding, competence and willingness to participate in care decisions have also been identified as associated with higher self-efficacy^57^. These two factors were not covered in this study.

## Conclusion

One-third of the participants have limited health literacy. Self-care was not associated with age, gender, CKD status, household income or education. Limited health literacy was associated with self-care, with strongest association with self-efficacy, followed by motivation and behaviour. More targeted approach can be adopted to improve self-care and health literacy amongst patients with CKD.

## Data Availability

Data cannot be shared freely due to local PDPA (Personal Data Protection Act) law. De-identified data can be made available to interested parties.

## Acknowledgement

The authors would like to thank the study participants, research assistants and staff at SingHealth Polyclinic Research Department for their support of this project.

